# Associations between regional adipose tissue distribution and skeletal muscle bioenergetics in older men and women

**DOI:** 10.1101/2023.11.10.23298359

**Authors:** Andrea M. Brennan, Paul M. Coen, Theresa Mau, Megan Hetherington-Rauth, Frederico G.S. Toledo, Erin E. Kershaw, Peggy M. Cawthon, Philip A. Kramer, Sofhia V. Ramos, Anne B. Newman, Steven R. Cummings, Daniel E. Forman, Reichelle X. Yeo, Giovanna DiStefano, Iva Miljkovic, Jamie N. Justice, Anthony J.A. Molina, Michael J. Jurczak, Lauren M. Sparks, Stephen B. Kritchevsky, Bret H. Goodpaster

## Abstract

**Objective:** Examine the association of ectopic adipose tissue (AT) with skeletal muscle (SM) mitochondrial bioenergetics in older adults.

**Methods:** Cross-sectional data from 829 older adults ≥70 years was used. Total abdominal, subcutaneous, and visceral AT; and thigh muscle fat infiltration (MFI) was quantified by MRI. SM mitochondrial energetics were characterized using *in vivo* ^31^P-MRS (ATP_max_) and *ex vivo* high-resolution respirometry (maximal oxidative phosphorylation (OXPHOS)). ActivPal was used to measure PA (step count). Linear regression models adjusted for covariates were applied, with sequential adjustment for BMI and PA.

**Results:** Independent of BMI, total abdominal (standardized (Std.) β=-0.21; R^2^=0.09) and visceral AT (Std. β=-0.16; R^2^=0.09) were associated with ATP_max_ (*p*<0.01), but not after further adjustment for PA (p≥0.05). Visceral AT (Std. β=-0.16; R^2^=0.25) and thigh MFI (Std. β=-0.11; R^2^=0.24) were negatively associated with carbohydrate-supported maximal OXPHOS independent of BMI and PA (*p*<0.05). Total abdominal AT (Std. β=-0.19; R^2^=0.24) and visceral AT (Std. β=-0.17; R^2^=0.24) were associated with fatty acid-supported maximal OXPHOS independent of BMI and PA (p<0.05).

**Conclusions:** Skeletal MFI and abdominal visceral, but not subcutaneous AT, are inversely associated with SM mitochondrial bioenergetics in older adults independent of BMI. Associations between ectopic AT and *in vivo* mitochondrial bioenergetics are attenuated by PA.

## Introduction

Age-related changes in body composition, including increases in total adiposity and concomitant decreases in lean body mass are strongly associated with metabolic dysfunction and risk for morbidity and mortality(1–3). Aging is also characterized by redistribution of adipose tissue (AT) from peripheral or subcutaneous regions to ectopic depots, including visceral and intermuscular AT(4, 5). Independent of total adiposity, accumulation of AT within these regions increases risk for several deleterious health outcomes including insulin resistance(6), cardiovascular disease(5), mobility limitations(7), and lower skeletal muscle (SM) strength and quality(8). Despite overwhelming evidence that accumulation of AT in ectopic fat depots has distinct functional and metabolic consequences, the molecular mechanisms underlying these associations are unclear.

Reductions in SM mitochondrial function and content are associated with many of the same harmful outcomes that occur with ectopic AT deposition, though whether excess ectopic AT is a cause, consequence, or correlate of SM mitochondrial function is unknown(9, 10). One of the primary functions of mitochondria is to produce adenosine triphosphate (ATP) through respiration or oxidative phosphorylation (OXPHOS), which subsequently fuels muscle contraction and movement in addition to several other fundamental cellular processes. Impaired mitochondrial metabolism is linked to age-associated risk for cardiometabolic disease and functional impairments, which can subsequently result in morbidity, mortality, and mobility disability in older adults(9). Reports in both middle-aged and older adults suggest that excess adiposity is associated with impaired SM mitochondrial metabolism(11–14). Muscle mitochondrial oxidative capacity and mitochondrial content are both negatively correlated with body mass index (BMI)(11). However, whether the accumulation of AT in ectopic and intermuscular depots, are associated with muscle mitochondrial energetics in older adults, independent of total adiposity, is not known.

Prior studies that examine region-specific associations between AT and SM mitochondrial activity are limited. Bharadwaj et al. studied citrate synthase activity and succinate-mediated respiration of isolated mitochondria from vastus lateralis biopsies, along with whole body and thigh composition from dual x-ray absorptiometry (DXA) and computed tomography (CT) in 25 healthy, sedentary, older men and women (>65 years of age)(11). They found that citrate synthase activity, but not succinate-mediated respiration, was negatively associated with total thigh fat volume, but not thigh intermuscular fat volume, independent of BMI and sex. In middle-aged adults, Bellissimo et al. observed associations between body composition measured by MRI and DXA and muscle mitochondrial function measured by phosphocreatine recovery time constant (τPCr) following standardized exercise in a MR scanner using ^31^P-magnetic resonance spectroscopy (^31^P-MRS) of the quadriceps muscle(15). Total fat mass, visceral AT, and thigh intermuscular AT were significantly and positively associated with τPCr (smaller values of τPCr reflect greater ATP synthesis). These studies suggest a link between reduced mitochondrial content and increased ectopic AT deposition, but are limited by relatively low sample sizes and incomplete measures of mitochondrial energetics and ectopic AT wherein all abdominal AT (subcutaneous and visceral) compartments were not reported. Furthermore, only one study(11) examined these parameters in older individuals, a population in which such outcomes are highly relevant to clinical and functional status.

Physical activity and energy expenditure also decrease with age(16), which can be related to both an increase in adiposity(17) and a decrease in SM mitochondrial energetics(18). It is possible that objectively measured physical activity may explain potential associations between regional AT depots and SM mitochondrial energetics in older adults.

The primary objective of this study was to examine associations between regional (abdominal and thigh) ectopic AT depots and SM mitochondrial energetics using both *in vivo* ^31^P-MRS and *ex vivo* high-resolution respirometry on permeabilized SM fibers from vastus lateralis in a large cohort of older men and women of the ongoing Study of Muscle, Mobility and Aging (SOMMA)(19). We hypothesized that increased ectopic (including both abdominal and intermuscular) AT will be associated with lower SM energetics in older adults, independent of BMI and physical activity.

## Methods

### Study cohort and recruitment

The Study of Muscle, Mobility and Aging (SOMMA) recruited men and women 70 years and older at the University of Pittsburgh and Wake Forest University School of Medicine from April 2019 to December 2021(19). Participants were eligible if they were able to walk 400 m at a usual pace and willing to undergo a muscle tissue biopsy and magnetic resonance (MR) scans. Those who appeared as they might not be able to complete the 400 m walk at the in-person screening visit completed a short distance walk (4 meters) to ensure their walking speed as ≥0.6m/s. Exclusion criteria included chronic anticoagulation therapy or other medical contraindication to biopsy or MR, inability to walk ¼ mile or climb a flight of stairs, BMI≥40 kg/m^2^ and active cancer or advanced chronic disease(19). This study was approved by the WIRB-Copernicus Group (WCG) Institutional Review Board (WCGIRB, study number 20180764). The number of participants at each step of the analysis is illustrated in **Figure 1**.

**Figure 1.**
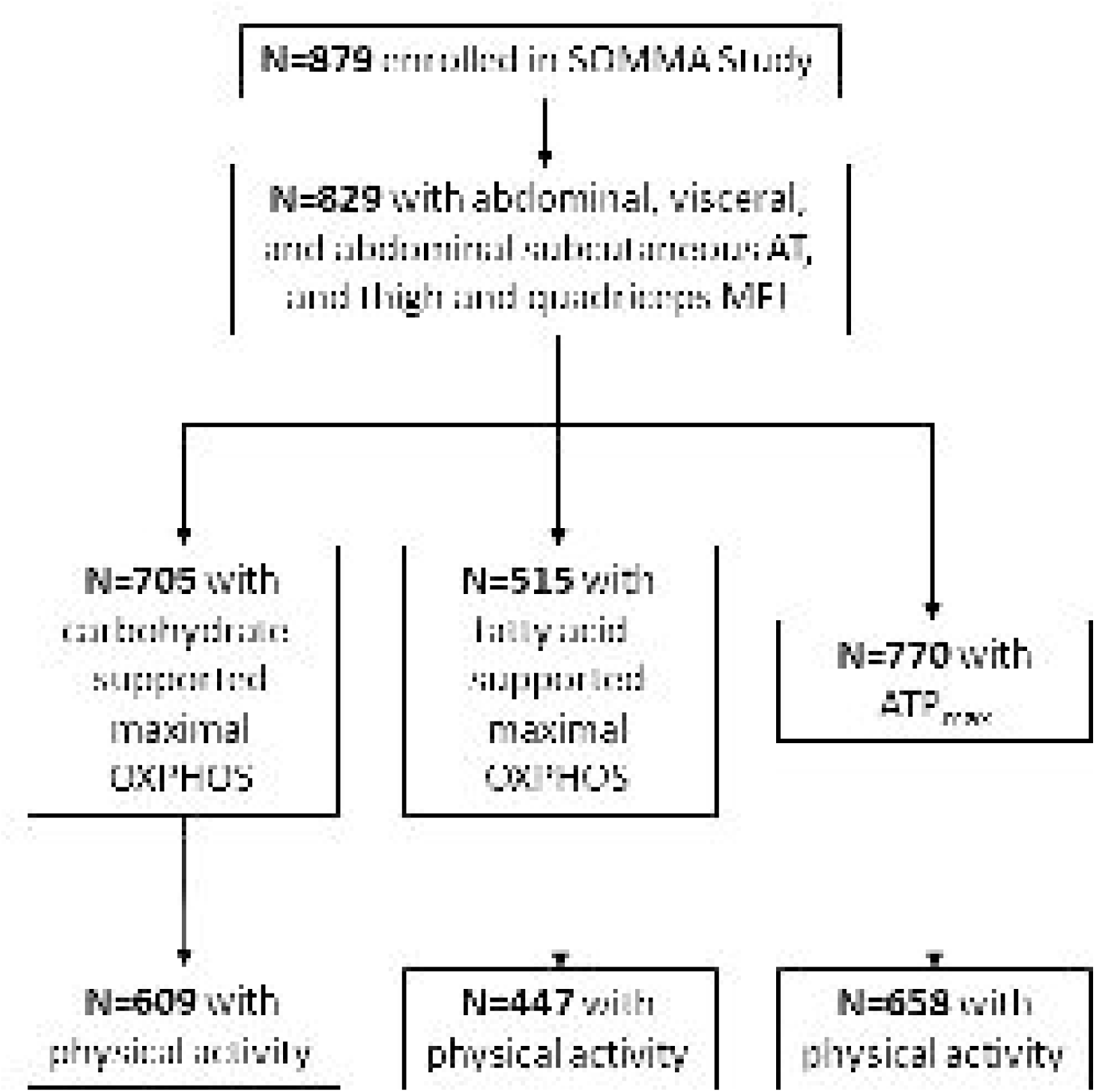
Participants selected for described statistical models. AT, adipose tissue; ATP_max_, maximal adenosine triphosphate production; OXPHOS, oxidative phosphorylation.

### Anthropometry

Height was measured using wall-mounted stadiometers and weight was measured via balance beams or digital scale. Waist circumference was also obtained.

### Body composition by MRI

A magnetic resonance imaging (MRI) scan was taken of the whole body to assess body composition. All MRI data was processed by the AMRA Company.

Images were analyzed using AMRA Researcher® (AMRA Medical AB, Linköping Sweden). Briefly, the image analysis consisted of image calibration, fusion of image stacks, image segmentation, and quantification of fat and muscle volumes(20, 21) and included manual quality control by a blinded trained operator. The following body composition parameters were obtained: abdominal subcutaneous AT, the subcutaneous AT volume in the abdomen from the top of the femoral head to the top of the thoracic vertebra T9; visceral AT, the AT volume within the abdominal cavity, excluding AT outside the abdominal skeletal muscles and AT within the cavity and posterior of the spine and back muscles; muscle fat infiltration (MFI), the mean fat fraction in the FFMV of the right and left anterior and posterior thigh. Total abdominal AT was calculated as the sum of abdominal subcutaneous and visceral AT volume.

### Muscle biopsy and fiber bundle processing

Percutaneous muscle biopsies of the vastus lateralis were conducted at the clinical sites using a Bergstrom trocar (5 or 6 mm) with suction as previously described(22, 23). Muscle tissue was dissected into fiber bundles and prepared for high-resolution respirometry using methods described in detail elsewhere(24).

### Ex vivo Mitochondrial respiration protocol

Following processing, permeabilized fiber bundles were transferred to the respiration chambers of an Oroboros O2k Oxygraph (Oroboros Inc., Innsbruck, Austria) in duplicate, with MiR05 supplemented with blebbistatin (25 μM) at 37°C. Oxygen levels were maintained between 400-200 μM. Two substrate protocols, carbohydrate (CHO)-supported and fatty acid (FA)-supported oxidative phosphorylation (OXPHOS), were run in parallel, each in duplicate. If muscle tissue and/or machine availability was limited, priority was given to CHO-supported OXPHOS.

For CHO-supported OXPHOS, maximal complex I&II-supported OXPHOS, referred to as CHO-supported max OXPHOS, was measured in the presence of pyruvate (5mM), malate (2mM), adenosine diphosphate (4.2mM), glutamate (10mM) and succinate (10mM); and carbonyl cyanide-p-trifluoromethoxyphenylhydrazone(FCCP) (1μM increments) was titrated to maximal oxygen consumption to measure maximal complex I– and II-supported electron transport system (ETS) capacity, referred to as max ETS.

For FA-supported OXPHOS, maximal complex I– and II-supported OXPHOS, referred to as FA-supported max OXPHOS, was measured in the presence of palmitoyl carnitine (25µM), malate (2mM), adenosine diphosphate (4.2mM), glutamate (10mM) and succinate (10mM).

All oxygen consumption measurements (pmol/s) were normalized to fiber bundle wet weight (pmol/s*mg), and data were analyzed in DatLab 7.4 software. Quality control measures included cytochrome c (10µM) injection to ensure mitochondrial outer membrane integrity, in which samples with a change >15% were omitted from the dataset. The mean coefficient of variation for duplicates of maximal OXPHOS measurement was 11.5% across both clinical sites(24).

### In Vivo ^31^P-MRS protocol

Maximal mitochondrial ATP production (ATPmax) following an acute bout of knee extensor exercise was determined *in vivo* using ^31^P magnetic resonance spectroscopy [MRS] to quantify the rate of phosphocreatine (PCr) recovery(25). The mean coefficient of variation for duplicates of ATP_max_ measurement was 9.9% across both clinic sites.

The exercise protocol was previously described(24). Briefly, participants laid supine with the right knee elevated at ∼30° in a 3.0 Tesla magnetic resonance magnet (Siemens Medical System – Prisma [Pittsburgh] or Skyra [Wake Forest]). Straps were placed over the legs and a 12′′ ^31^P/^1^H dual – tuned, surface RF coil (PulseTeq, Limited) was placed over the quadriceps. Sixty seconds after commencing acquisition of 76 sequential spectra, participants kicked repeatedly as fast as they could for 30 seconds. After rapid processing of data to determine breakdown and acidosis, a second or third data series was acquired adjusting exercise duration to satisfy breakdown and acidosis criteria. The protocol was designed to deplete PCr stores by 30% – 50% to ensure high signal to noise defining PCr recovery without inducing acidosis (pH <6.8), which inhibits oxidative phosphorylation. PCr recovery rate after exercise until PCr returned to baseline levels was fit and the time-constant of the monoexponential fit (tau) was used to calculate maximum mitochondrial ATP production (ATPmax)(26). All data were analyzed using jMRUI v6.0(27).

### ActivPal Activity Monitoring

Physical activity was measured as step count (steps per day) using the ActivPal Activity monitor by PAL Technologies Ltd (Glasgow, Scotland). Participants wore the monitor on the midline of the right thigh for 7 consecutive 24-hour periods, including during bathing or water sports. The ActivPal device contains an accelerometer that records at 20Hz and proprietary algorithms are used to determine step count through thigh position and acceleration. The algorithm for determining a valid day used a 24-hour wear period and allowed for up to 4 hours of non-wear per day. Raw. datx files using PALconnect were processed using PALanalysis (version V8.11.1.49).

### Statistical Analyses

A linear test for trend following one-way ANOVAs were used to examine differences in baseline characteristics between tertiles of abdominal AT. Pearson’s correlation (*r*) coefficients were used to assess bivariate associations between muscle mitochondrial energetics and AT depots, age, BMI, and physical activity. Multiple linear regression models were used to estimate associations between AT depots and muscle mitochondrial energetics, adjusted for relevant covariates in 3 models. Model 1 adjusted for site (*in vivo* mitochondrial energetics) or technician (*ex vivo* mitochondrial energetics), race (White or non-White), age, and sex. Model 2 adjusted for Model 1 plus BMI. Model 3 adjusted for Model 2 plus physical activity. Each AT depot was assessed individually; thus, multiple AT depots were not included in the same model. An interaction term for sex and each AT depot was assessed for all models. Standardized betas (Std. β; measured in units of standard deviation), model fit (R^2^) and p-values were reported. Statistical analysis and visualization were completed using JMP version 16.1.0 (SAS Institute Inc.), and Graphpad Prism version 9.3.1 for Windows (GraphPad Software, San Diego, California, USA) software packages.

## Results

### Participant Characteristics

Participant characteristics are reported by tertiles of total abdominal AT in **Table 1**. Apart from sex, there were significant linear trends for baseline variables across abdominal AT tertiles (all p<0.05).

**Table 1.**
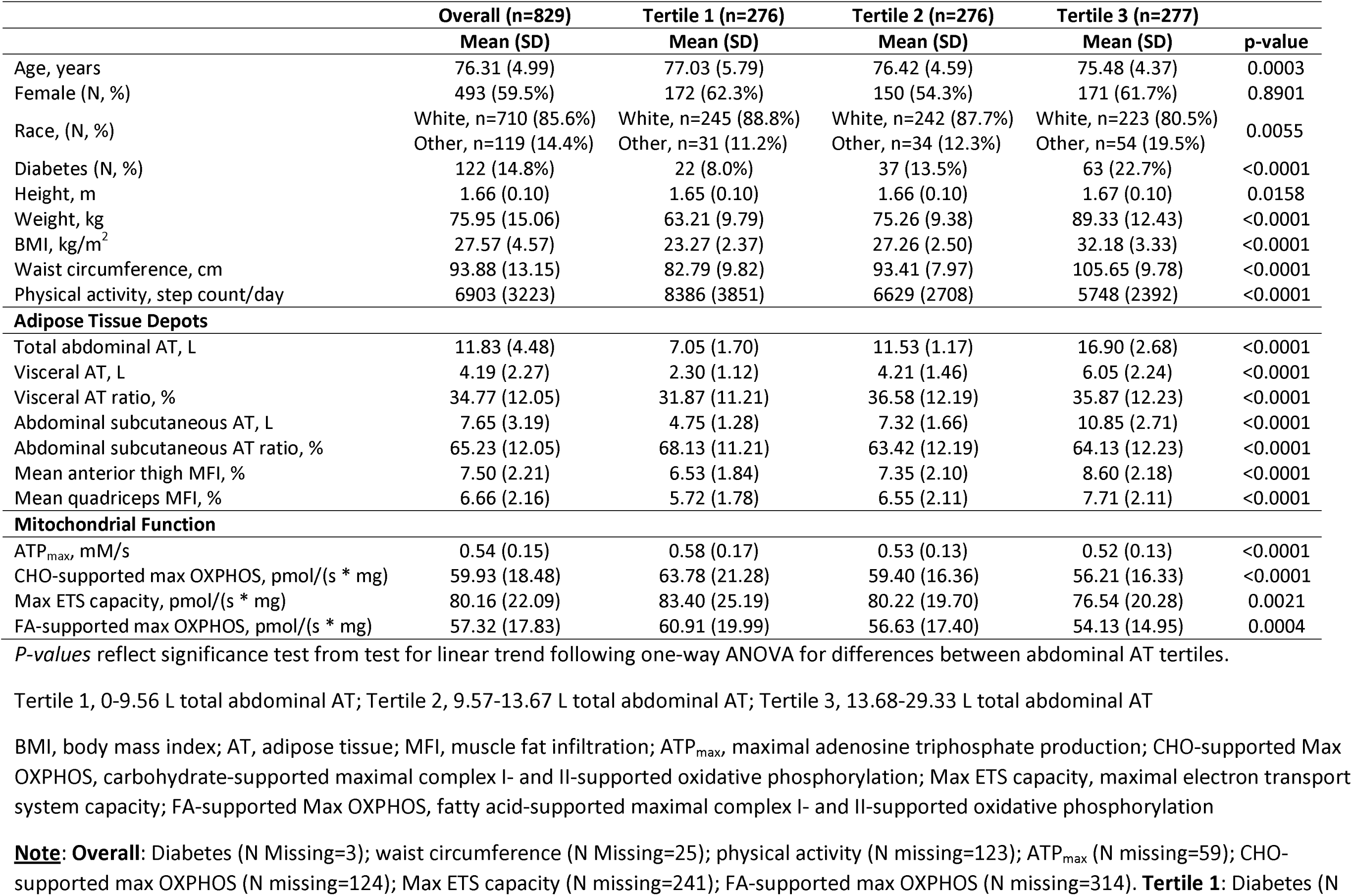
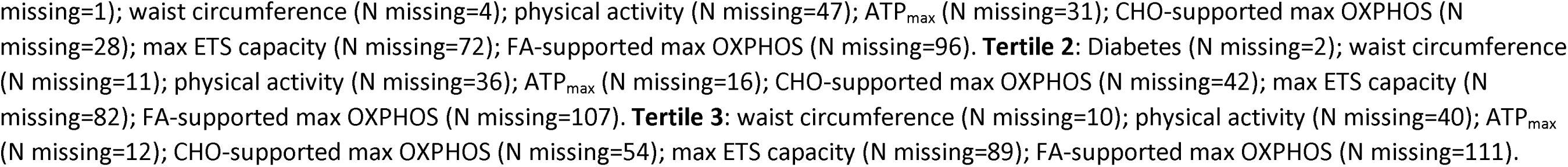
Participant Baseline Characteristics by Tertiles of Total Abdominal Adipose Tissue: The Study of Muscle, Mobility and Aging.

|  | Overall (n=829) | Tertile 1 (n=276) | Tertile 2 (n=276) | Tertile 3 (n=277) |  |
| --- | --- | --- | --- | --- | --- |
|  | Mean (SD) | Mean (SD) | Mean (SD) | Mean (SD) | p-value |
| Age, years | 76.31 (4.99) | 77.03 (5.79) | 76.42 (4.59) | 75.48 (4.37) | 0.0003 |
| Female (N, %) | 493 (59.5%) | 172 (62.3%) | 150 (54.3%) | 171 (61.7%) | 0.8901 |
| Race, (N, %) | White, n=710 (85.6%) | White, n=245 (88.8%) | White, n=242 (87.7%) | White, n=223 (80.5%) | 0.0055 |
|  | Other, n=119 (14.4%) | Other, n=31 (11.2%) | Other, n=34 (12.3%) | Other, n=54 (19.5%) |  |
| Diabetes (N, %) | 122 (14.8%) | 22 (8.0%) | 37 (13.5%) | 63 (22.7%) | <0.0001 |
| Height, m | 1.66 (0.10) | 1.65 (0.10) | 1.66 (0.10) | 1.67 (0.10) | 0.0158 |
| Weight, kg | 75.95 (15.06) | 63.21 (9.79) | 75.26 (9.38) | 89.33 (12.43) | <0.0001 |
| BMI, kg/m<sup>2</sup> | 27.57 (4.57) | 23.27 (2.37) | 27.26 (2.50) | 32.18 (3.33) | <0.0001 |
| Waist circumference, cm | 93.88 (13.15) | 82.79 (9.82) | 93.41 (7.97) | 105.65 (9.78) | <0.0001 |
| Physical activity, step count/day | 6903 (3223) | 8386 (3851) | 6629 (2708) | 5748 (2392) | <0.0001 |
| **Adipose Tissue Depots** |  |  |  |  |  |
| Total abdominal AT, L | 11.83 (4.48) | 7.05 (1.70) | 11.53 (1.17) | 16.90 (2.68) | <0.0001 |
| Visceral AT, L | 4.19 (2.27) | 2.30 (1.12) | 4.21 (1.46) | 6.05 (2.24) | <0.0001 |
| Visceral AT ratio, % | 34.77 (12.05) | 31.87 (11.21) | 36.58 (12.19) | 35.87 (12.23) | <0.0001 |
| Abdominal subcutaneous AT, L | 7.65 (3.19) | 4.75 (1.28) | 7.32 (1.66) | 10.85 (2.71) | <0.0001 |
| Abdominal subcutaneous AT ratio, % | 65.23 (12.05) | 68.13 (11.21) | 63.42 (12.19) | 64.13 (12.23) | <0.0001 |
| Mean anterior thigh MFI, % | 7.50 (2.21) | 6.53 (1.84) | 7.35 (2.10) | 8.60 (2.18) | <0.0001 |
| Mean quadriceps MFI, % | 6.66 (2.16) | 5.72 (1.78) | 6.55 (2.11) | 7.71 (2.11) | <0.0001 |
| **Mitochondrial Function** |  |  |  |  |  |
| ATP<sub>max</sub>, mM/s | 0.54 (0.15) | 0.58 (0.17) | 0.53 (0.13) | 0.52 (0.13) | <0.0001 |
| CHO-supported max OXPHOS, pmol/(s * mg) | 59.93 (18.48) | 63.78 (21.28) | 59.40 (16.36) | 56.21 (16.33) | <0.0001 |
| Max ETS capacity, pmol/(s * mg) | 80.16 (22.09) | 83.40 (25.19) | 80.22 (19.70) | 76.54 (20.28) | 0.0021 |
| FA-supported max OXPHOS, pmol/(s * mg) | 57.32 (17.83) | 60.91 (19.99) | 56.63 (17.40) | 54.13 (14.95) | 0.0004 |

*P-values* reflect significance test from test for linear trend following one-way ANOVA for differences between abdominal AT tertiles.
Tertile 1, 0-9.56 L total abdominal AT; Tertile 2, 9.57-13.67 L total abdominal AT; Tertile 3, 13.68-29.33 L total abdominal AT
BMI, body mass index; AT, adipose tissue; MFI, muscle fat infiltration; ATP<sub>max</sub>, maximal adenosine triphosphate production; CHO-supported Max OXPHOS, carbohydrate-supported maximal complex I- and II-supported oxidative phosphorylation; Max ETS capacity, maximal electron transport system capacity; FA-supported Max OXPHOS, fatty acid-supported maximal complex I- and II-supported oxidative phosphorylation
**Note:** **Overall:** Diabetes (N Missing=3); waist circumference (N Missing=25); physical activity (N missing=123); ATP<sub>max</sub> (N missing=59); CHO-supported max OXPHOS (N missing=124); Max ETS capacity (N missing=241); FA-supported max OXPHOS (N missing=314). **Tertile 1:** Diabetes (N
missing=1); waist circumference (N missing=4); physical activity (N missing=47); ATP<sub>max</sub> (N missing=31); CHO-supported max OXPHOS (N missing=28); max ETS capacity (N missing=72); FA-supported max OXPHOS (N missing=96). **Tertile 2:** Diabetes (N missing=2); waist circumference (N missing=11); physical activity (N missing=36); ATP<sub>max</sub> (N missing=16); CHO-supported max OXPHOS (N missing=42); max ETS capacity (N missing=82); FA-supported max OXPHOS (N missing=107). **Tertile 3:** waist circumference (N missing=10); physical activity (N missing=40); ATP<sub>max</sub> (N missing=12); CHO-supported max OXPHOS (N missing=54); max ETS capacity (N missing=89); FA-supported max OXPHOS (N missing=111).

### Bivariate Correlations of Muscle Mitochondrial Energetics, Adipose Tissue, Age, BMI, and Physical Activity

Pearson correlations of all continuous variables are shown in **Figure 2**. Older age was associated with lower muscle mitochondrial energetics (ATP_max_, CHO– and FA-supported max OXPHOS), total abdominal and abdominal subcutaneous AT, BMI, and physical activity, but higher thigh and quadriceps MFI (all p<0.01). Higher physical activity was associated with higher muscle mitochondrial energetics (ATP_max_, CHO– and FA-supported max OXPHOS, max ETS) and lower adiposity (abdominal subcutaneous AT, visceral AT, thigh MFI, and quadriceps MFI) (all p<0.0001). ATP_max_, CHO-supported max OXPHOS, FA-supported max OXPHOS, and max ETS were negatively associated with all AT depots (all p<0.01) except for visceral AT (p≥0.05).

**Figure 2.**
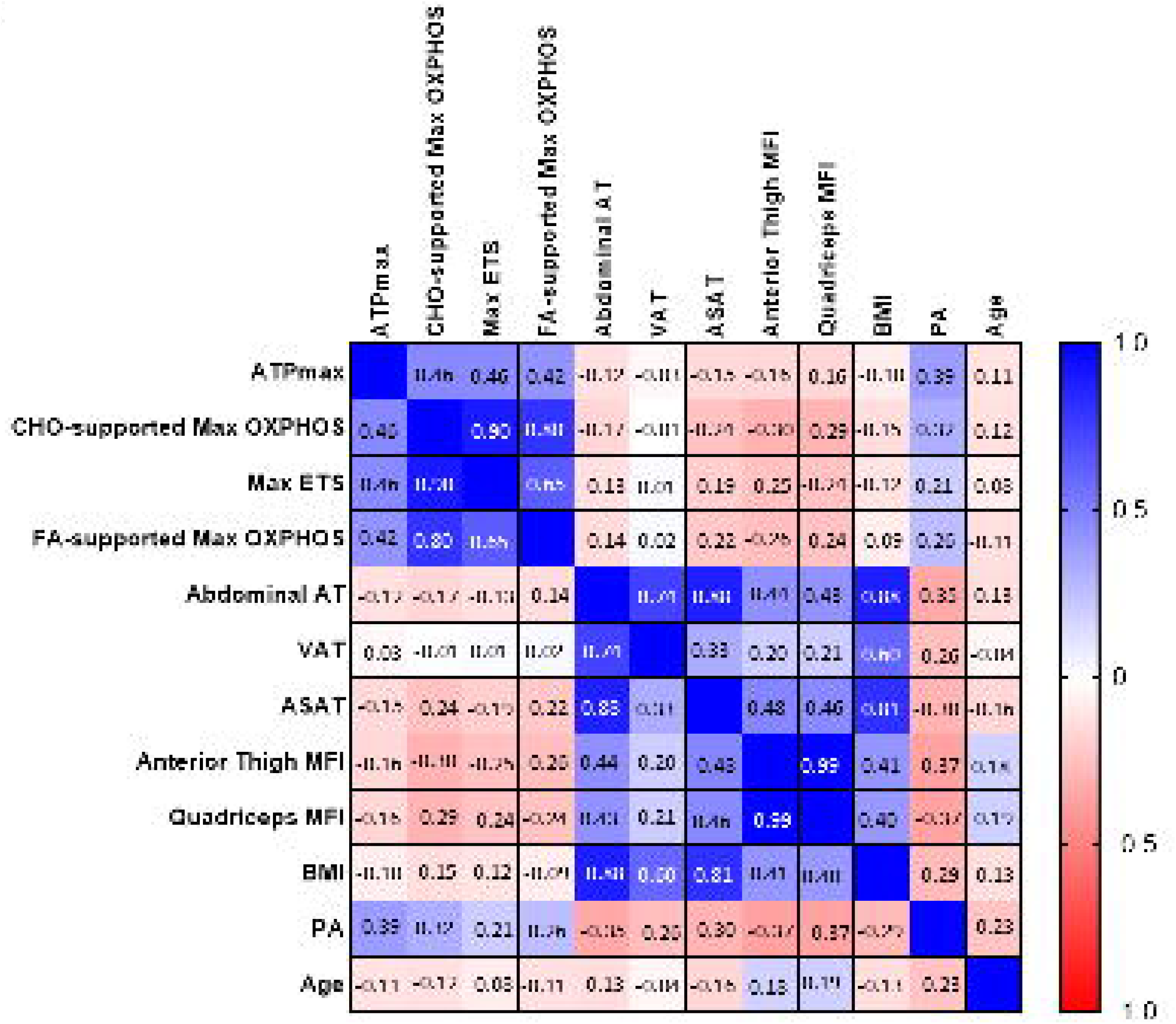
Bivariate correlation matrix of skeletal muscle mitochondrial energetics, continuous covariates, and adipose tissue depots. Pearson’s correlation (*r*) displayed for each pair of variables. Increasing blue intensity reflects correlations closer to +1.0 while increasing shades of red represent correlations closer to –1.0. White squares indicate a non-significant correlation (*p*≥0.05). ATPmax, maximal adenosine triphosphate production; CHO, carbohydrate; Max, maximal; OXPHOS, oxidative phosphorylation; FA, fatty acid; ETS, electron transport system capacity; AT, adipose tissue; VAT, visceral AT; ASAT, abdominal subcutaneous AT; MFI, muscle fat infiltration; BMI, body mass index; PA, physical activity.

### In vivo mitochondrial function (ATPmax)

Linear regression models for the relationships between AT depots and ATP_max_ are summarized in **Table 2**. Among all participants, total abdominal AT (Std. β=-0.13), visceral AT (Std. β=-0.16), abdominal subcutaneous AT (Std. β=-0.12), anterior thigh MFI (Std. β=-0.09) and quadriceps MFI (Std. β=-0.10) were significantly negatively associated with ATP_max_ after adjusting for sex, site, race, and age (all p<0.05). The addition of BMI in the model eliminated these associations, except for total abdominal AT (Std. β=-0.21) and visceral AT (Std. β=-0.16) which remained negatively associated with ATP_max_ (p<0.05). After adjusting for physical activity, there were no significant associations between adipose tissue depots and ATP_max_. The low R^2^ in Model 1, ranging from 0.08-0.09 remains relatively unchanged in Model 2 after adjusting for BMI. However, after further adjusting for physical activity, the R^2^ increased to 0.20 suggesting that physical activity explains more of the variance in ATP_max_ than total adiposity reflected by BMI.

**Table 2.** Associations of adipose tissue (AT) depots and ^31^P-MRS derived ATP (mM/s).

|  | **Model 1**<br>(sex, site, race, and age) |  |  | **Model 2**<br>(Model 1 + BMI) |  |  | **Model 3**<br>(Model 2 + physical activity) |  |  |
| --- | --- | --- | --- | --- | --- | --- | --- | --- | --- |
|  | n | Std. β | Model Fit (R<sup>2</sup>) | n | Std. β | R<sup>2</sup> | n | Std. β | R<sup>2</sup> |
| Abdominal AT | 770 | -0.13** | 0.09 | 770 | -0.21** | 0.09 | 658 | -0.06 | 0.20 |
| Abdominal Subcutaneous AT | 770 | -0.12** | 0.08 | 770 | -0.10 | 0.08 | 658 | 0.01 | 0.20 |
| VAT | 770 | -0.16** | 0.09 | 770 | -0.16** | 0.09 | 658 | -0.08 | 0.20 |
| Anterior Thigh MFI | 770 | -0.09* | 0.08 | 770 | -0.06 | 0.08 | 658 | 0.04 | 0.20 |
| Quadriceps MFI | 770 | -0.10** | 0.08 | 770 | -0.07 | 0.08 | 658 | 0.02 | 0.20 |

\*p<0.05; \*\*p<0.01; \*\*\*p<0.0001
<sup>31</sup>P-MRS, <sup>31</sup>phosphorus magnetic resonance spectroscopy; ATPmax, maximal adenosine triphosphate production; AT, adipose tissue; ASAT, abdominal subcutaneous AT; VAT, visceral AT; MFI, muscle fat infiltration Standardized beta (Std. β) measured in units of standard deviation.

### Ex vivo muscle mitochondrial energetics

Linear regression models for the relationship between AT depots and CHO-supported max OXPHOS are summarized in **Table 3**. Total abdominal AT (Std. β=-0.15), abdominal subcutaneous AT (Std. β=-0.13), visceral AT (Std. β=-0.18), anterior thigh MFI (Std. β=-0.18) and quadriceps MFI (Std. β=-0.17) were negatively associated with CHO-supported max OXPHOS, adjusted for sex, technician, race and age (p<0.01). After adding BMI to the model, total abdominal AT (Std. β=-0.17), visceral AT (Std. β=-0.15), anterior thigh MFI (Std. β=-0.14) and quadriceps MFI (Std. β=-0.14) remained significantly associated (p<0.05). Visceral AT (Std. β=-0.16), anterior thigh (Std. β=-0.11) and quadriceps MFI (Std. β=-0.10) remained significantly associated with CHO-supported max OXPHOS after further adjusting for physical activity (p<0.05). Associations between max ETS and AT depots mirrored those with CHO-supported max OXPHOS, shown in **Table 4**, with the exception that both total abdominal AT (Std. β=-0.19) and visceral AT (Std. β=-0.20), in addition to anterior thigh MFI (Std. β=-0.14) and quadriceps MFI (Std. β=-0.13) remained significantly associated with max ETS after adjusting for BMI and physical activity.

**Table 3.** Associations of adipose tissue (AT) depots and CHO-supported max OXPHOS (pmol/(s * mg)).

|  | **Model 1**<br>(sex, tech, race, and age) |  |  | **Model 2**<br>(Model 1 + BMI) |  |  | **Model 3**<br>(Model 2 + physical activity) |  |  |
| --- | --- | --- | --- | --- | --- | --- | --- | --- | --- |
| | n | Std. $\beta$ | Model Fit ($R^2$) | n | Std. $\beta$ (SD) | $R^2$ | n | Std. $\beta$ | $R^2$ |
| Abdominal AT | 705 | -0.15*** | 0.20 | 705 | -0.17* | 0.20 | 609 | -0.15 | 0.24 |
| Abdominal Subcutaneous AT | 705 | -0.13** | 0.19 | 705 | -0.06 | 0.19 | 609 | -0.02 | 0.24 |
| VAT | 705 | -0.18*** | 0.20 | 705 | -0.15** | 0.20 | 609 | -0.16** | 0.25 |
| Anterior Thigh MFI | 705 | -0.18*** | 0.20 | 705 | -0.14** | 0.20 | 609 | -0.11* | 0.24 |
| Quadriceps MFI | 705 | -0.17*** | 0.20 | 705 | -0.14** | 0.20 | 609 | -0.10* | 0.24 |

\* $p < 0.05$ ; \*\* $p < 0.01$ ; \*\*\* $p < 0.0001$
CHO, carbohydrate; max OXPHOS, maximal oxidative phosphorylation; AT, adipose tissue; ASAT, abdominal subcutaneous AT; VAT, visceral AT; MFI, muscle fat infiltration
Standardized beta (Std. $\beta$ ) measured in units of standard deviation.

**Table 4.** Associations of adipose tissue (AT) depots and maximal capacity of the electron transport system (pmol/(s * mg)).

|  | **Model 1**<br>(sex, tech, race, and age) |  |  | **Model 2**<br>(Model 1 + BMI) |  |  | **Model 3**<br>(Model 2 + physical activity) |  |  |
| --- | --- | --- | --- | --- | --- | --- | --- | --- | --- |
| | n | Std. $\beta$ | Model Fit ($R^2$) | n | Std. $\beta$ | $R^2$ | n | Std. $\beta$ | $R^2$ |
| Abdominal AT | 588 | -0.15** | 0.13 | 588 | -0.16 | 0.13 | 497 | -0.19* | 0.16 |
| Abdominal Subcutaneous AT | 588 | -0.13** | 0.12 | 588 | -0.04 | 0.12 | 497 | -0.02 | 0.16 |
| VAT | 588 | -0.17** | 0.13 | 588 | -0.14* | 0.13 | 497 | -0.20** | 0.17 |
| Anterior Thigh MFI | 588 | -0.18*** | 0.14 | 588 | -0.15** | 0.14 | 497 | -0.14** | 0.17 |
| Quadriceps MFI | 588 | -0.18*** | 0.14 | 588 | -0.15** | 0.14 | 497 | -0.13** | 0.17 |

\*p<0.05; \*\*p<0.01; \*\*\*p<0.0001
AT, adipose tissue; ASAT, abdominal subcutaneous AT; VAT, visceral AT; MFI, muscle fat infiltration
Standardized beta (Std. $\beta$ ) measured in units of standard deviation.

Linear regression models for the relationship between AT depots and FA-supported max OXPHOS are summarized in **Table 5**. Abdominal AT (Std. β=-0.12), abdominal subcutaneous AT (Std. β=-0.10), visceral AT (Std. β=-0.14), anterior thigh MFI (Std. β=-0.10), and quadriceps MFI (Std. β=-0.09) were significantly associated with FA-supported max OXPHOS adjusted for sex, technician, race, and age (p<0.05). After adding BMI and PA to the model, total abdominal AT (Std. β=-0.19) and visceral AT (Std. β=-0.17) remained significantly associated with FA-supported max OXPHOS (p<0.05).

**Table 5.** Associations of adipose tissue (AT) depots and FA-supported max OXPHOS (pmol/(s * mg)).

|  | **Model 1**<br>(sex, tech, race, and age) |  |  | **Model 2**<br>(Model 1 + BMI) |  |  | **Model 3**<br>(Model 2 + physical activity) |  |  |
| --- | --- | --- | --- | --- | --- | --- | --- | --- | --- |
| | n | Std. $\beta$ | Model Fit ($R^2$) | n | Std. $\beta$ | $R^2$ | n | Std. $\beta$ | $R^2$ |
| Abdominal AT | 515 | -0.12** | 0.23 | 515 | -0.23** | 0.23 | 447 | -0.19* | 0.24 |
| Abdominal Subcutaneous AT | 515 | -0.10* | 0.22 | 515 | -0.12 | 0.22 | 447 | -0.05 | 0.23 |
| VAT | 515 | -0.14** | 0.23 | 515 | -0.16* | 0.23 | 447 | -0.17* | 0.24 |
| Anterior Thigh MFI | 515 | -0.10* | 0.22 | 515 | -0.09 | 0.22 | 447 | -0.04 | 0.24 |
| Quadriceps MFI | 515 | -0.09* | 0.22 | 515 | -0.07 | 0.22 | 447 | -0.03 | 0.23 |

\*p<0.05; \*\*p<0.01; \*\*\*p<0.0001
FA, fatty acid; max OXPHOS, maximal oxidative phosphorylation; AT, adipose tissue; ASAT, abdominal subcutaneous AT; VAT, visceral AT; MFI, muscle fat infiltration
Standardized beta (Std. $\beta$ ) measured in units of standard deviation.

Except for FA-supported max OXPHOS, in which the sex interaction term for total abdominal and abdominal subcutaneous AT was significant (p<0.05), the sex interaction term for all AT depots and CHO-supported max OXPHOS, max ETS, and ATP_max_ was not significant in any model.

## Discussion

The primary finding of this study is that regional adiposity, particularly abdominal visceral AT and SM fat infiltration is associated with SM mitochondrial bioenergetics in older adults, although these associations with *in vivo* measures are significantly mitigated by BMI and habitual physical activity. Considering all confounders, SM mitochondrial bioenergetics measured *in vivo* was not significantly correlated with AT depots. Even without physical activity in the model, total abdominal and visceral AT accounted for only a minimal amount of the variance (8-9%) in SM bioenergetics, implying nominal impact of regional adiposity above and beyond obesity. For *ex vivo* mitochondrial bioenergetics measures, thigh AT and visceral AT correlated with carbohydrate-supported mitochondrial energetics, while total abdominal AT and visceral AT correlated with fatty acid-supported SM mitochondrial bioenergetics, independent of BMI and physical activity.

Various features of SM mitochondrial function, including oxidative and phosphorylation activity(9, 28, 29) as well as morphological changes(30) have been consistently shown to decline with age(9). Similarly, individuals with obesity have reduced levels of oxidative enzymes, lower mitochondrial content, decreased activity of the electron transport chain, and reduced oxygen consumption compared to lean individuals(31–36). However, many of these studies assessed the relationship of obesity and muscle mitochondrial energetics in young to middle-aged individuals, typically with more advanced stages of obesity. Although the inter-relationships between mitochondrial energetics, aging, and obesity have been reported(11), albeit in a limited number of studies with small sample sizes, whether alterations in mitochondrial function is a cause or consequence of obesity or vice versa is unknown. In addition, it is possible that SM mitochondrial energetics and obesity are not directly related to each other but are manifestations of a common underlying mechanism. Excess adiposity, both total and regional can release proinflammatory cytokines and hormones that over time may diminish muscle mitochondrial capacity(37); however, dysregulation of fuel utilization by impaired mitochondria may disrupt whole-body energy balance and promote both generalized and ectopic obesity and its related comorbidities(38, 39).

Although there is evidence for a relationship between generalized obesity in young/middle aged adults and SM mitochondrial function, the impact of regional AT distribution on mitochondrial energetics is unclear, especially in older adults. Aging is characterized by a shift in AT distribution, wherein AT accumulates in ectopic regions, such as SM and the visceral compartment of the abdomen(4). Our findings suggest that the relationship between ectopic AT and mitochondrial energetics differs by energy substrate and whether mitochondrial energetics are measured *ex vivo* or *in vivo*. After adjusting for BMI both visceral AT and thigh MFI was inversely associated with carbohydrate-supported maximal OXPHOS. In contrast, only total abdominal AT and visceral AT were inversely associated with fatty acid-supported OXPHOS. These *ex vivo* findings differ from those of Bharadwaj et al., who reported that no regional depots were associated with succinate-mediated respiration. The authors, however, did not measure abdominal AT depots, and measured thigh AT via computed tomography which may explain the discrepancy.(11)

*In vivo* maximal ATP_max_ was inversely associated with total abdominal and visceral AT but not thigh MFI after adjusting for BMI. Though *in vivo* ^31^P MRS measurement of ATPmax is complementary to high-resolution respirometry in permeabilized muscle fibers, there are distinct and inherent methodological differences that may explain the discordant findings. ^31^P MRS measures maximal ATP synthesis following muscle contraction *in vivo* which is influenced by the cellular environment including substrate supply, oxygen saturation, calcium signaling and intracellular pH(28, 40). In contrast, high-resolution respirometry in permeabilized muscle fibers from biopsies assesses mitochondrial energetics under highly controlled conditions(41). It is possible that *ex vivo* high-resolution respirometry more directly and robustly describes the relationship between mitochondrial energetics and AT distribution, without influence from physiological factors that cannot be controlled with *in vivo* measures. It is also possible that the source and supply of substrate, i.e., fatty acids or carbohydrates to fuel mitochondrial energetics *in vivo* impacts the associations with ectopic AT, where *ex vivo* high resolution respirometry substrates are saturating.

Physical activity, through structured exercise or non-exercise activity can impact fat distribution and mitochondrial energetics. After adjusting for physical activity, there were no significant associations between AT depots and *in vivo* SM mitochondrial energetics. Indeed, both visceral AT and thigh MFI remained significantly associated with carbohydrate-supported maximal OXPHOS, but only visceral AT was correlated with fatty acid-supported maximal OXPHOS. Interestingly, the only difference between the two *ex vivo* protocols at maximal OXPHOS is the absence of fatty acids in the carbohydrate-supported protocol and the absence of pyruvate in the fatty acid-supported protocol. Taken together, our findings suggest that physical activity may ameliorate damaging effects of ectopic thigh AT by increasing fatty acid oxidation. This is supported by others who have shown that exercise induces fatty acid oxidation in the muscle(42) and prevents accumulation of toxic lipid intermediates(43). Furthermore, our findings are also supported by prior studies which show that physical activity and not weight loss *per se* is required for improvements in mitochondrial function(44, 45). This observation is encouraging, as it implies that the potential harmful consequences on mitochondrial function incurred by aging-related ectopic AT accumulation may be mitigated by increased physical activity in older adults. However, the independent association between visceral AT and thigh MFI and *ex vivo* carbohydrate-supported SM mitochondrial energetics supports the need for further investigation into underlying mechanisms and potential targets for therapeutic intervention.

There are strengths and limitations in this study. While abdominal and SM AT were measured, additional ectopic fat depots that may be closely linked to metabolic impairments, including fat deposits in the legs and thigh outside of the muscle, and pancreatic and cardiac AT, could not be assessed. Our observations are cross-sectional, precluding the ability to determine a directional causal relationship between ectopic AT and mitochondrial energetics. Aging-associated changes in AT distribution and its influence on age-related impairments in muscle bioenergetics will be important to understand in longitudinal studies. The majority of participants identified as non-Hispanic White, so generalizability to other race and ethnicities may be limited. A key strength of this study is that these measures of muscle mitochondria and regional adiposity were performed in a large group of older women and men, providing a more robust analysis of the relationship between muscle mitochondrial energetics and body composition in later life. Another notable strength is the use of state-of-the-art assessments of both *in vivo* and *ex vivo* mitochondrial function and AT distribution, increasing the quality, accuracy and validity of the measurements.

## Conclusion

Aging-associated impairments in mitochondrial capacity and accumulation of AT in ectopic depots are both linked to the pathogenesis of metabolic and functional disorders(31, 38). Overall, our findings suggest associations between regional AT depots and SM mitochondrial energetics that are attenuated after adjusting for BMI and physical activity. With exception, visceral AT and fatty acid-supported maximal OXPHOS, and both visceral AT and thigh MFI and carbohydrate-supported maximal OXPHOS, remained associated after adjusting for confounders. The range in total variance explained between separate AT depots within each model is minimal, indicating that the AT depot per se does not contribute significantly to the total model variance. Longitudinal and mechanistic studies are required to determine directionality of the associations between adiposity and SM mitochondrial energetics to support development of interventions that target improved metabolic health with aging.

## Data Availability

All data produced in the present study are available upon reasonable request to the authors.

## Acknowledgements

For a full list of personnel who contributed to the SOMMA study, please see “Cummings SR, Newman AB, Coen PM, Hepple RT, Collins R, Kennedy K, Danielson M, Peters K, Blackwell T, Johnson E, Mau T, Shankland EG, Lui LY, Patel S, Young D, Glynn NW, Strotmeyer ES, Esser KA, Marcinek DJ, Goodpaster BH, Kritchevsky S, Cawthon PM. The Study of Muscle, Mobility and Aging (SOMMA). A Unique Cohort Study about the Cellular Biology of Aging and Age-related Loss of Mobility. J Gerontol A Biol Sci Med Sci. 2023 Feb 9:glad052. doi: 10.1093/gerona/glad052. Epub ahead of print. PMID: 36754371.”

